# Development of Cas13a-based Assays for *Neisseria gonorrhoeae* Detection and Gyrase A Determination

**DOI:** 10.1101/2023.05.21.23290304

**Authors:** Lao-Tzu Allan-Blitz, Palak Shah, Gordon Adams, John A. Branda, Jeffrey D. Klausner, Robert Goldstein, Pardis C. Sabeti, Jacob E. Lemieux

## Abstract

**Background:** *Neisseria gonorrhoeae* is one of the most common bacterial sexually transmitted infections. The emergence of antimicrobial-resistant *N. gonorrhoeae* is an urgent public health threat. Currently, diagnosis of *N. gonorrhoeae* infection requires expensive laboratory infrastructure, while antimicrobial susceptibility determination requires bacterial culture, both of which are infeasible in low-resource areas where prevalence is highest. Recent advances in molecular diagnostics, such as Specific High-sensitivity Enzymatic Reporter unLOCKing (SHERLOCK) using CRISPR-Cas13a and isothermal amplification, have the potential to provide low-cost detection of pathogen and antimicrobial resistance.

**Methods and Results:** We designed and optimized RNA guides and primer-sets for SHERLOCK assays capable of detecting *N. gonorrhoeae* via the *por*A gene and of predicting ciprofloxacin susceptibility via a single mutation in the gyrase A (*gyr*A) gene. We evaluated their performance using both synthetic DNA and purified *N. gonorrhoeae* isolates. For *por*A, we created both a fluorescence-based assay and lateral flow assay using a biotinylated FAM reporter. Both methods demonstrated sensitive detection of 14 *N. gonorrhoeae* isolates and no cross-reactivity with 3 non-gonococcal *Neisseria* isolates. For *gyr*A, we created a fluorescence-based assay that correctly distinguished between 20 purified *N. gonorrhoeae* isolates with phenotypic ciprofloxacin resistance and 3 with phenotypic susceptibility. We confirmed the *gyr*A genotype predictions from the fluorescence-based assay with DNA sequencing, which showed 100% concordance for the isolates studied.

**Conclusion:** We report the development of Cas13a-based SHERLOCK assays that detect *N. gonorrhoeae* and differentiate ciprofloxacin-resistant isolates from ciprofloxacin-susceptible isolates.

## Introduction

*Neisseria (N.) gonorrhoeae* is one of the most common bacterial sexually transmitted infections worldwide (1). There were an estimated 87 million cases reported in 2016 (1), with the highest prevalence among low-resource settings (2–4), which is likely to be an underestimate due to under-reporting. The consequences of inadequately treated infection can be serious, ranging from pelvic inflammatory disease (5), infertility (6), and neonatal blindness (7), to an increased risk for HIV infection (8–13).

Furthermore, antimicrobial resistance in *N. gonorrhoeae* is a global public health threat (14, 15). *N. gonorrhoeae* has developed resistance to nearly all antimicrobials used in its treatment (16). Because culture is not routinely performed and standard-of care nucleic acid amplification testing (NAAT) via polymerase chain reaction (PCR) does not provide information on antibiotic susceptibility, all *N. gonorrhoeae* infections in the United States are treated with third generation cephalosporins, further driving selective pressure towards the emergence of resistance (16, 17). Recent reports of resistance to third generation cephalosporins (18–22) have raised concern for untreatable infection. In response, the United States Centers for Disease Control and Prevention has increased the recommended dose of ceftriaxone for treating gonorrhea (23). However, treatment of *N. gonorrhoeae* infection with antibiotics no longer empirically recommended due to high levels of resistance has been made possible by rapid molecular assays detecting genotypic markers of resistance (16, 17, 24). Use of such assays might reduce the spread of cephalosporin resistance (25).

Neither PCR for pathogen detection, nor bacterial culture for susceptibility determination are available in low-resource settings, as PCR requires expensive laboratory infrastructure and culture can be laborious and time-intensive for *N.* gonorrhoeae (26). Consequently, treatment of *N. gonorrhoeae* infection is limited to syndromic management in low-resource settings, which is insensitive for case finding (27–30) and further drives the emergence of antimicrobial resistance (16, 17). In fact, limited data suggest that low-resource areas have some of the highest prevalence of antimicrobial-resistant *N. gonorrhoeae* infections (31–33). Thus, the World Health Organization’s action plan for combating the emergence of antimicrobial resistance calls for the development of rapid molecular assays for pathogen detection and predicting antimicrobial susceptibility (34). Previous work has indicated that the *por*A gene may be a useful target for *N. gonorrhoeae* detection (35), and that phenotypic resistance to ciprofloxacin is predicted by the presence of a single nucleotide polymorphism at codon 91 of the gyrase A (*gyr*A) gene (36, 37). Such testing, however, still requires PCR capabilities, which are generally inaccessible in low-resource settings.

Specific high-sensitivity enzymatic reporter unlocking (SHERLOCK) technology utilizes Cas13a, a CRISPR enzyme paired with isothermal amplification via recombinase polymerase amplification (RPA) (38, 39), a low-cost, sensitive, specific, and field-deployable diagnostic technology (40, 41). Cas13a-based detection works via complementary binding of programmable CRISPR guide RNA (gRNA) sequences to target sequences, which activates the inherent Cas13a-meated collateral cleavage of an RNA reporter (38, 42). Such assays can be employed with standard fluorescence reports or adapted for paper-based lateral flow detection (43). Moreover, Cas13a has been shown to have high specificity with reduced tolerance for activation with increasing mismatches between gRNA and the template, which can facilitate discriminating between strains containing point mutations. In this study, we aimed to develop SHERLOCK assays for *N. gonorrhoeae* detection and *gyr*A genotype determination. We explored fluorescence-based and lateral flow readouts for each assay, and evaluated their performance using *N. gonorrhoeae* synthetic DNA and purified isolates. We aimed for this work to be a first step towards developing methods for *N. gonorrhoeae* detection and antimicrobial resistance determination accessible anywhere in the world.

## Methods

### Ethics statement

The Mass General Brigham Institutional Review Board approved this study under protocols 2019P003305 and 2020P000323.

### Reagents and Materials

Detailed information on reagents used and stock concentrations can be found in Supplemental Tables 1 and 2.

### Synthetic DNA preparation and DNA extraction from purified isolates

We tested assays were using both synthetic *N. gonorrhoeae* DNA and purified *N. gonorrhoeae* isolates. We prepared synthetic DNA samples by serial dilution from commercially purchased (Integrated DNA Technologies, United States) double-stranded DNA (dsDNA) of the *gyr*A target region into water. We stored purified isolates in glycerol at –80° C prior to extraction. We extracted whole-genomic DNA from *N. gonorrhoeae* purified isolates using the DNeasy Blood and Tissue Kit (Qiagen, Germany). The starting volume for extraction was 400 μL, and extracted DNA was eluted into 100 μL of nuclease-free water. With each isolate, we were provided minimum inhibitory concentrations (MICs) in µg/mL for ciprofloxacin, obtained using standard methods. Additionally, we purchased non-gonococcal *Neisseria* isolates from American Type Culture Collection (ATCC) and cultured by the Massachusetts General Clinical Microbiology Laboratory: *N. meningitidis* (ATCC 13077)*, perflava* (ATCC 14799), and *lactamica* (ATCC 23970). The performance of the *por*A assay was also assessed on those isolates.

We quantified the concentration of extracted *N. gonorrhoeae* DNA using qPCR. The forward and reverse primer sequences for the *N. gonorrhoeae gyr*A gene were 5’ GCGACGGCCTAAAGCCAGTG 3’ and 5’ GTCTGCCAGCATTTCATGTGAG 3’, respectively. Those primers were provided by a previous study (44). The qPCR reaction mixtures contained 1× FastStart SYBR Green Master Mix (Sigma Aldrich, United States), 0.5 µM of each primer, and DNA template in a 1:9 template to master mix ratio. We adjusted the final qPCR reaction volume to 10 µL with nuclease-free water, and loaded in triplicate on a 384-well plate, which was run on a QuantStudio 6 (Applied Biosystems, United States) with the following cycle conditions: heat activation at 95° C for 3 minutes, 40 cycles of a denaturing step at 95° C for 15 seconds, an annealing step at 60° C for 1 minute, and an extension step at 72° C, followed by a final extension step at 68° C for 2 minutes. We collected amplification data during the second extension stage and analyzed those data using the Standard Curve module of the Applied Biosystems Analysis Software. We quantified clinical Isolates against a standard curve, which showed an average concentration of 1,000 copies per mL across isolates. Subsequently, we evaluated thermal DNA extraction by resuspending 3 purified isolates in 100 µL of water and heating the isolates to 95°C for 10 minutes in accordance with prior protocols (45).

### Guide RNA and primer design for N. gonorrhoeae detection

Cas13a gRNAs have two components: the fixed “handle” region to which the Cas13a protein binds, and a 28-nucleotide “spacer” region complementary to the target. The nucleotide sequence of the spacer can be chosen by the user to confer the specificity of the assay. We selected the *por*A gene of *N. gonorrhoeae* for pathogen detection as has been used previously (35). We used an online software package ADAPT (Activity-Informed Design with All-Inclusive Patrolling of Targets; https://adapt.run) (46), which applies an algorithm for optimal Cas13a gRNAs design, was used for guide design, and selected 3 gRNAs from the output of that software targeting different locations in the *por*A gene.

We designed forward and reverse RPA primers using National Center for Biotechnology Information (NCBI) Primer-Basic Local Alignment Search Tool (BLAST), which were synthesized by Integrated DNA Technologies (United States). We developed 2 primer sets per guide location (total of six primer sets), which were 27 to 35 nucleotides in length. The primer-sets had melting temperatures between 58° C and 68° C, and produced amplicons of 140 to 200 base pairs in length. We appended a T7 RNA polymerase promoter sequence (5’ GAAATTAATACGACTCACTATAGG 3’) to the 5’ end of the forward primers of each set to allow for T7 transcription.

### One-pot SHERLOCK assay

We performed SHERLOCK reactions using 45 nM C2c2 *Lwa*Cas13a (GenScript Biotech Corp, United States) resuspended in 1× storage buffer (SB: 50 mM Tris (pH 7.5), 600 mM KCl, 5% glycerol, and 2mM dithiothreitol (DTT)) such that the resuspended protein was at 2.25 μM, 1 U/μL murine RNase Inhibitor (NEB), 10 U/μL T7 RNA polymerase (Lucigen), 136 nM RNaseAlert substrate v2 (ThermoFisher Scientific, United States), 1× SHINE Buffer (SHINE: 20 mM HEPES (4-(2-hydroxyethyl)-1– piperazineethanesulfonic acid) (pH 8.0), 60 mM KCl, and 5% polyethylene glycol (PEG)), and 2 mM of each rNTP (NEB).

We rehydrated the TwistAmp Basic Kit lyophilized pellets (1 pellet per 73.42 μL master mix volume) using the prepared master mix. We added 14 mM MgAOc (TwistDx, United Kingdom) after resuspension to activate the RPA pellets. We then subdivided the master mix for each guide-primer set pair being analyzed, to which was added 22.5 nM gRNA (Integrated DNA Technologies, United States) and 320 nM each of the RPA primers (Integrated DNA Technologies, United States). We prepared SHERLOCK reactions to 70 μL and loaded as 20 μL triplicates into a 384-well plate, with a ratio of 1:5 master mix to sample. We measured fluorescence by the BioTek Cytation 5 plate reader (BioTek, United States) over 3 hours at 37 °C, with readings every 5 minutes (excitation: 485; emission: 528) for quantitative detection.

### Lateral Flow Detection

To convert to lateral flow readout, we modified the SHERLOCK master mix to exchange substrate v2 for a biotinylated FAM reporter at a final concentration of 1 µM. We incubated samples at 37 °C for 90 minutes per existing protocols to allow for optimal RPA amplification. Following incubation, we added 80 μL HybriDetect assay buffer (Milennia Biotec, Germany) to each sample in a 1:5 dilution along with a HybriDetect lateral flow strip (Milennia Biotec, Germany). We inspected strips and took images using a smartphone camera 3-5 minutes after the strips were added.

### Confirmatory DNA Sequencing

We performed whole-genome sequencing on extracted DNA samples following the Illumina DNA Prep manufacturer protocol (Illumina, United States). We constructed and pooled libraries using the Illumina DNA Prep Kit. We measured library concentrations on a Qubit4 Flourometer using the Qubit High Sensitivity 1× dsDNA kit (ThermoFisher Scientific, United States), while we measured the average library size on an Agilent TapeStation 4150 using the Agilent High Sensitivity D1000 ScreenTape kit (Agilent Technologies, United States). We conducted genomic sequencing on an Illumina MiniSeq instrument (Illumina, United States).

### Data Analysis

We subtracted baseline fluorescence (at 0 min) from fluorescence values through reaction progression. We averaged the final ten fluorescence values of each replicate to provide the reported fluorescence values. We compared mean differences in fluorescence using Student’s t test, with significance defined as p<0.05. We interpreated lateral flow readouts by visual inspection. We generated all figures in PRISM Software Version 9.5.1 (GraphPad, United States).

## Results

### N. gonorrhoeae detection via a Cas13a-based porA assay

To create an assay for *N. gonorrhoeae* detection, we first designed 6 *por*A primer-guide pairs and evaluated their performance, both in terms of high sensitivity and low cross-reactivity, using a fluorescence-based readout (Figure 1). We performed initial testing on 3 purified *N. gonorrhoeae* isolates using both negative template controls as well as synthetic *gyr*A as a positive control. We selected guide 2, primer set 2 as it produced both a high fluorescent signal and excellent discrimination between synthetic *N. gonorrhoeae* purified isolates and the negative controls. We excluded guide 3 primer set 1 due to cross-reactivity with the *gyr*A control.

**Figure 1:**
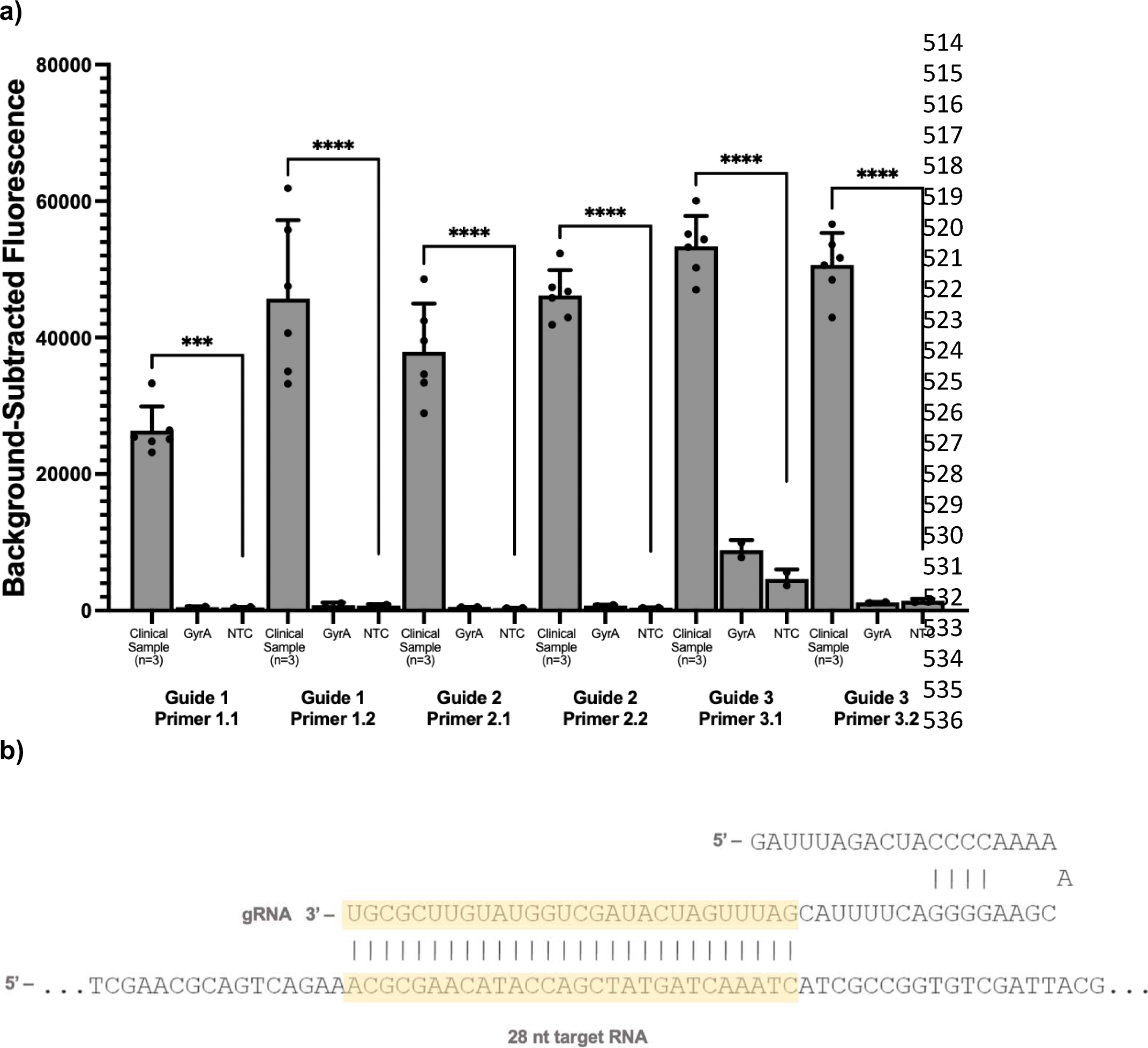
Guide and Primer Selection for a Cas13a-based assay for detecting *N. gonorrhoeae*. Figure 1 Legend: Panel a) shows the performance of three guides targeting different regions of the *por*A gene tested on 3 *N. gonorrhoeae* purified isolates as well as synthetic *gyr*A template as a control and a negative template control (NTC). Panel b) shows the selected *por*A guide sequence. *** Indicates statistically significant differences in florescence at the *p<0.05* level

Having selected our gRNA and primer-set for *por*A detection, we evaluated the limit of detection (LoD) using serial dilutions in water as well as the detection of purified *N. gonorrhoeae* isolates using a fluorescence-based readout. The *por*A assay had a LoD of 10,000 copies per mL (Figure 2a). We then tested the assay on 14 purified isolates and 3 non-gonococcal *Neisseria* isolates: *N. meningitidis, N. perflava,* and *N. lactamica*. The assay detected all 14 *N. gonorrhoeae* isolates, with peak fluorescence occurring after 20 minutes, and did not detect any of the non-gonococcal *Neisseria* isolates.

**Figure 2:**
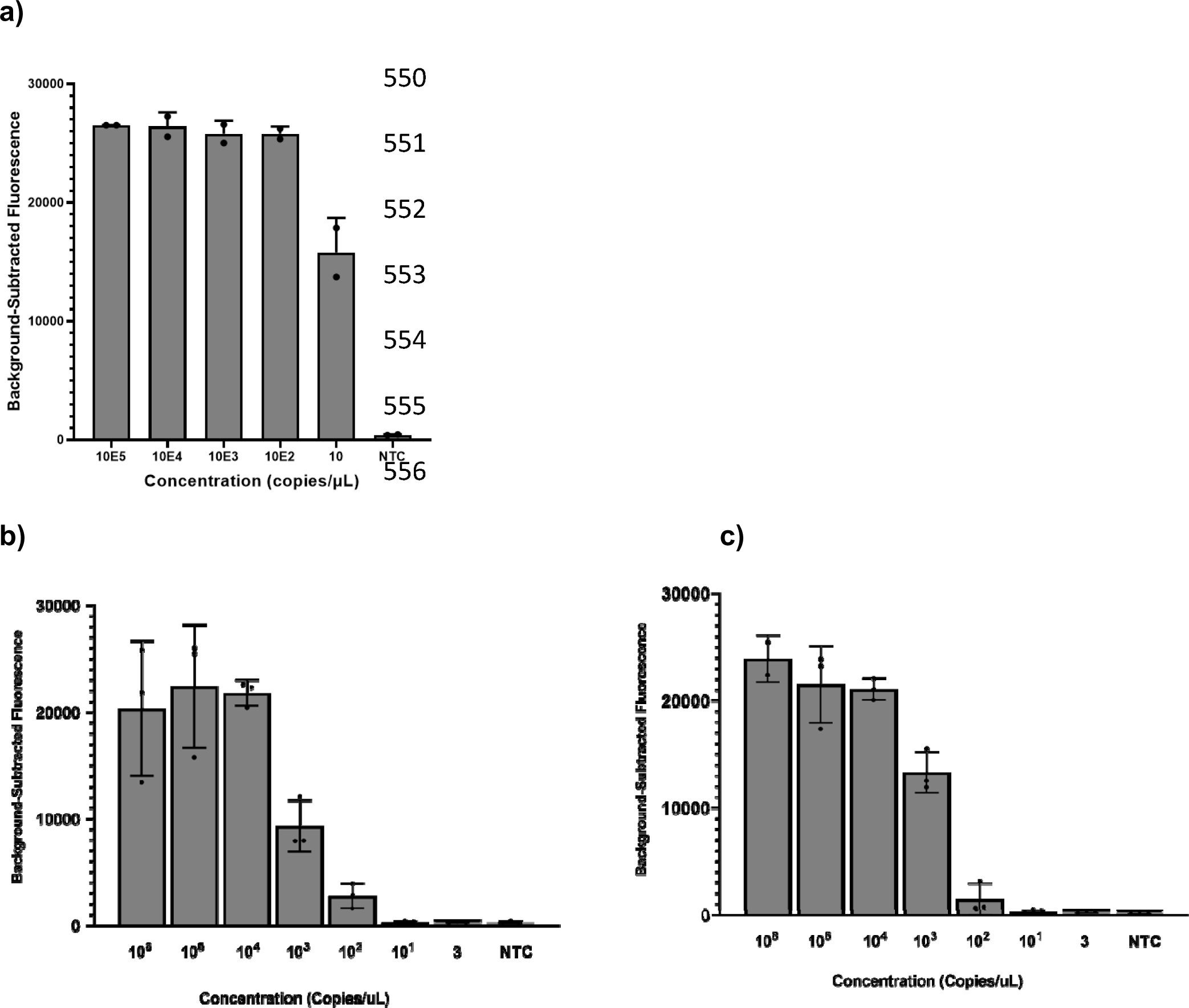
I*n vitro* limit of detection of the Cas13a *N. gonorrhoeae* and *gyr*A genotypic assays. Figure 2 Legend: Panel a) shows the limit of detection of the *N. gonorrhoeae* Cas13a detection assay using the selected guide-primer set for the *por*A gene among purified *N. gonorrhoeae* isolates and a negative template control (NTC); Panel b) shows the limit of detection of the Cas13a-based assay using the wildtype guide against synthetic wildtype DNA target; Panel c) shows the limit of detection of Cas13a-based assay using the mutant guide against synthetic mutant DNA target. The serial dilutions of synthetic DNA were done in water.

We then assessed the *N. gonorrhoeae por*A detection assay using a lateral flow readout, substituting the standard fluorescence reporter with a biotinylated FAM reporter compatible with the test strips. Based on prior protocols, we allocated 90 minutes for the assay. Visual inspection of the test strips 3-5 minutes after specimen introduction revealed detection of all 14 purified isolates tested in triplicate (Figure 3a) and excellent discrimination between *N. gonorrhoeae* and 3 non-gonococcal *Neisseria* isolates (Figure 3b).

**Figure 3:**
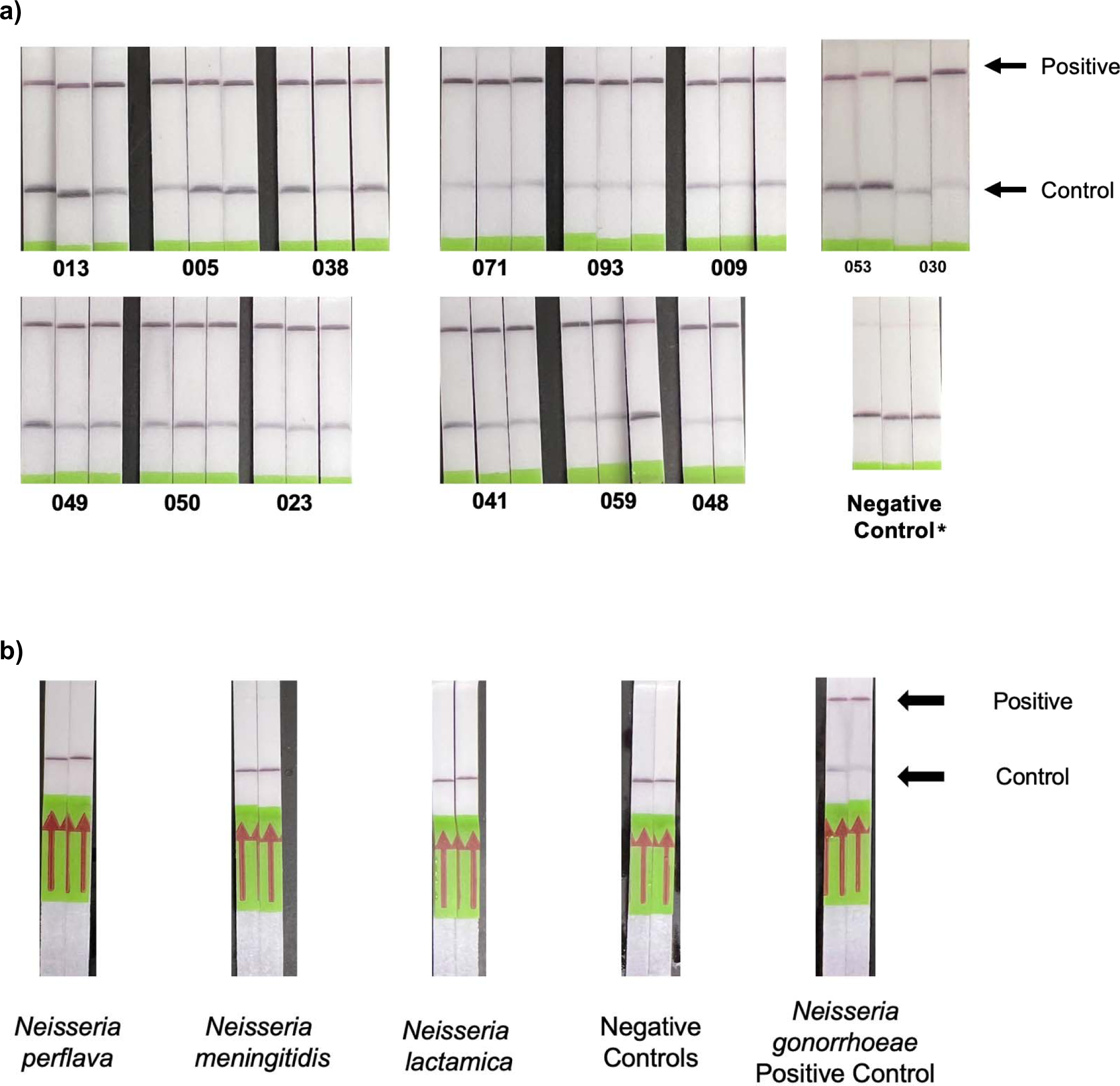
Performance of a Cas13a-Based Lateral Flow Assay for Detecting *N. gonorrhoeae*. Figure 3 Legend: Panel a) shows the performance of the Cas13a-based lateral flow assay on 14 purified *N. gonorrhoeae* isolates tested in triplicate. Panel b) shows the discrimination of the lateral flow assay for *N. gonorrhoeae* isolates compared with non– gonococcal *Neisseria* isolates. * The faint band at the test line in the negative control is expected per the manufacturer protocol

Having shown that we can develop a lateral flow-based *N. gonorrhoeae* detection assay, we explored the possibility of simplifying upstream DNA extraction to facilitate deployment in low-resource settings. To do so, we evaluated fluorescence *N. gonorrhoeae* detection on 3 purified isolates that underwent thermal DNA extraction. All 3 isolates were detected using the selected guide primer set combination, and showed excellent amplification.

### GyrA genotype determination via a Cas13a-based assay

To create an assay for predicting *N. gonorrhoeae* resistance to ciprofloxacin, we first designed 2 guide pairs (wildtype and mutant) to target the point mutation in codon 91 of the *gyr*A gene and three flanking primer sets. We placed the mutation of interest 3 nucleotides distal to the Cas hairpin, previously shown to be the optimal position (47). We placed an additional synthetic mismatch in either the 2^nd^ position or the 4^th^ position of the spacer region. We elected to design the guides manually instead of using ADAPT given the precise mutation of interest was known. Placing the synthetic mutation at the 2^nd^ position produced the highest fluorescence and greatest discrimination between the wildtype and mutant synthetic DNA targets (Supplemental Figure 1). We tested three forward and reverse primer sets for use with that guide, and selected the set that produced the highest fluorescence signal and greatest discrimination between the wildtype and mutant synthetic DNA targets (Supplemental Figure 2). We evaluated the *in vitro* LoD of the fluorescence-based *gyr*A assay via serial dilutions in water of synthetic wildtype and mutant DNA targets. The *gyr*A assay had a LoD of 1,000,000 copies per mL for both wildtype and mutant targets (Figure 2b).

To further assess the performance of the *gyr*A assay, we analyzed 23 purified *N. gonorrhoeae* isolates with susceptibility to ciprofloxacin determined phenotypically by culture and genotypically by sequencing to detect mutation codon 91 of the *gyr*A gene. We used a MIC breakpoint of ≥ 1 µg/mL to define ciprofloxacin resistance (Table 1). Of the 23 isolates, 20 with MICs between 1 and >16 µg/mL were deemed resistant, and 3 with MICs all < 0.015 µg/mL were deemed susceptible. Of the 20 *N. gonorrhoeae* isolates with MICs ≥ 1 µg/mL, 100% had mutant *gyr*A genotypes by DNA sequencing. Of the 3 *N. gonorrhoeae* isolates with MICs < 0.015 µg/mL, 100% had no mutation at codon 91 of the *gyr*A gene by DNA sequencing. Supplemental Figure 3 shows the phylogenetic tree of the 23 *N. gonorrhoeae* isolates, demonstrating that the assay works reliably across phylogenetically diverse isolates.

**Table 1:**
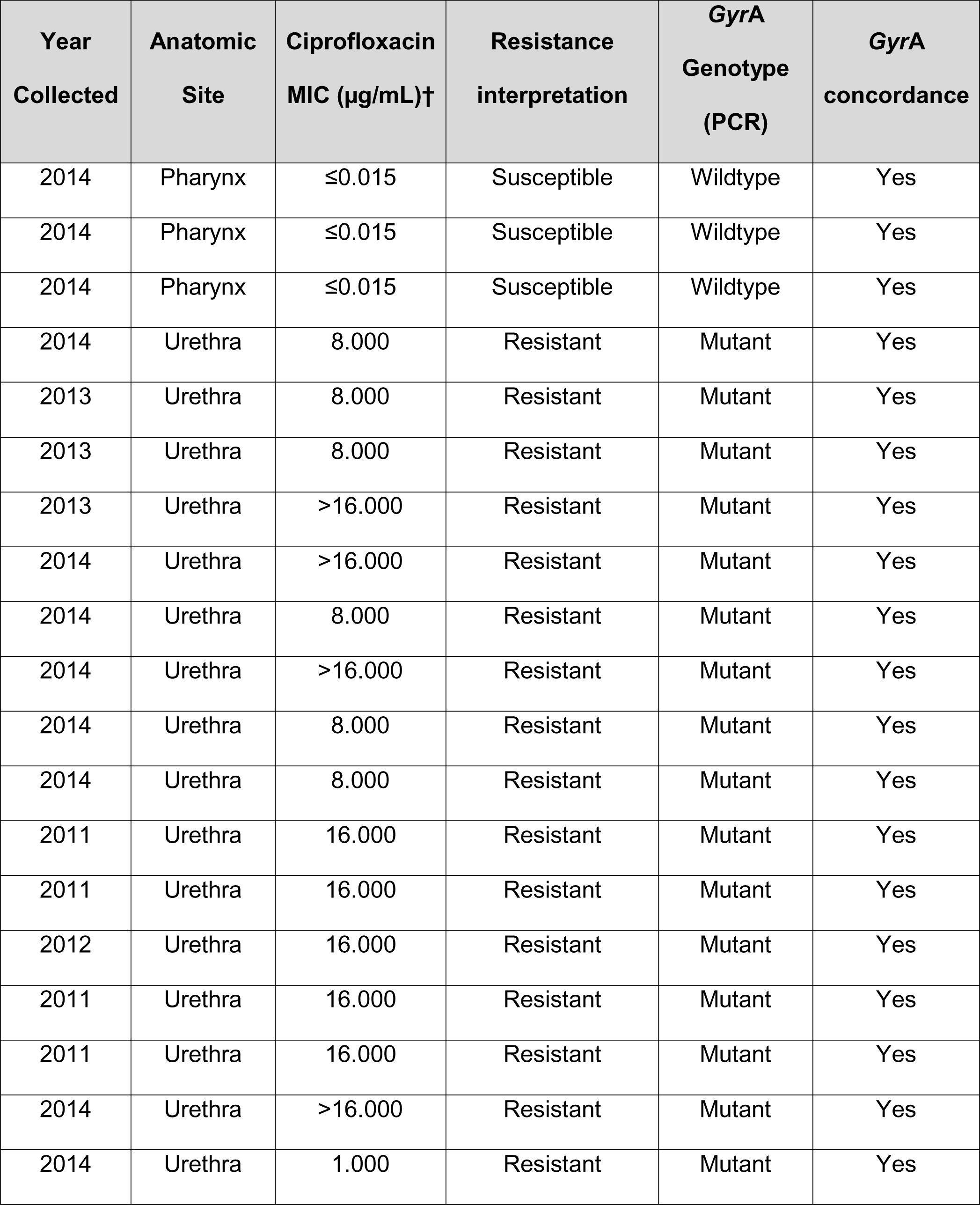
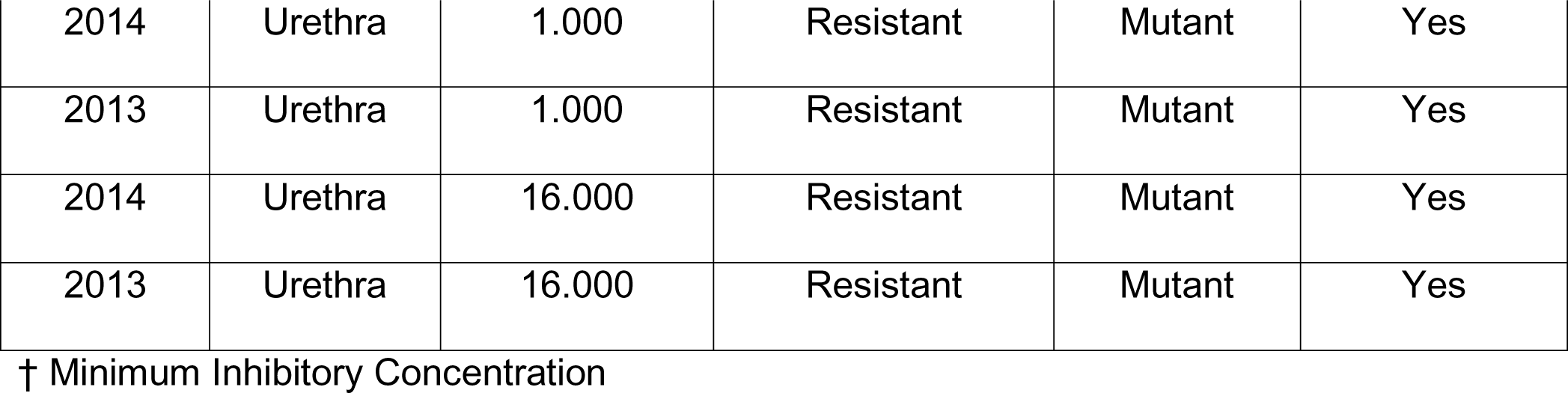
Characteristics of purified *N. gonorrhoeae* isolates

| **Year Collected** | **Anatomic Site** | **Ciprofloxacin MIC (µg/mL)†** | **Resistance interpretation** | **GyrA Genotype (PCR)** | **GyrA concordance** |
| --- | --- | --- | --- | --- | --- |
| 2014 | Pharynx | ≤0.015 | Susceptible | Wildtype | Yes |
| 2014 | Pharynx | ≤0.015 | Susceptible | Wildtype | Yes |
| 2014 | Pharynx | ≤0.015 | Susceptible | Wildtype | Yes |
| 2014 | Urethra | 8.000 | Resistant | Mutant | Yes |
| 2013 | Urethra | 8.000 | Resistant | Mutant | Yes |
| 2013 | Urethra | 8.000 | Resistant | Mutant | Yes |
| 2013 | Urethra | >16.000 | Resistant | Mutant | Yes |
| 2014 | Urethra | >16.000 | Resistant | Mutant | Yes |
| 2014 | Urethra | 8.000 | Resistant | Mutant | Yes |
| 2014 | Urethra | >16.000 | Resistant | Mutant | Yes |
| 2014 | Urethra | 8.000 | Resistant | Mutant | Yes |
| 2014 | Urethra | 8.000 | Resistant | Mutant | Yes |
| 2011 | Urethra | 16.000 | Resistant | Mutant | Yes |
| 2011 | Urethra | 16.000 | Resistant | Mutant | Yes |
| 2012 | Urethra | 16.000 | Resistant | Mutant | Yes |
| 2011 | Urethra | 16.000 | Resistant | Mutant | Yes |
| 2011 | Urethra | 16.000 | Resistant | Mutant | Yes |
| 2014 | Urethra | >16.000 | Resistant | Mutant | Yes |
| 2014 | Urethra | 1.000 | Resistant | Mutant | Yes |
| 2014 | Urethra | 1.000 | Resistant | Mutant | Yes |
| 2013 | Urethra | 1.000 | Resistant | Mutant | Yes |
| 2014 | Urethra | 16.000 | Resistant | Mutant | Yes |
| 2013 | Urethra | 16.000 | Resistant | Mutant | Yes |

† Minimum Inhibitory Concentration

We evaluated the discrimination of the selected wildtype and mutant guides for codon 91 of the *gyr*A gene among all 23 isolates. All of the 20 ciprofloxacin-resistant *gyr*A mutant specimens were detected by the mutant Cas13a assay, while 0 of the 3 wildtype isolates were detected by the mutant Cas13a assay, showing a 100% agreement. Figure 4 shows the pooled performance among all specimens, while Supplemental Figure 4 shows the performance on each individual specimen. Figure 5 shows the DNA sequence alignment for all 23 isolates with the wildtype and mutant RNA guides.

**Figure 4:**
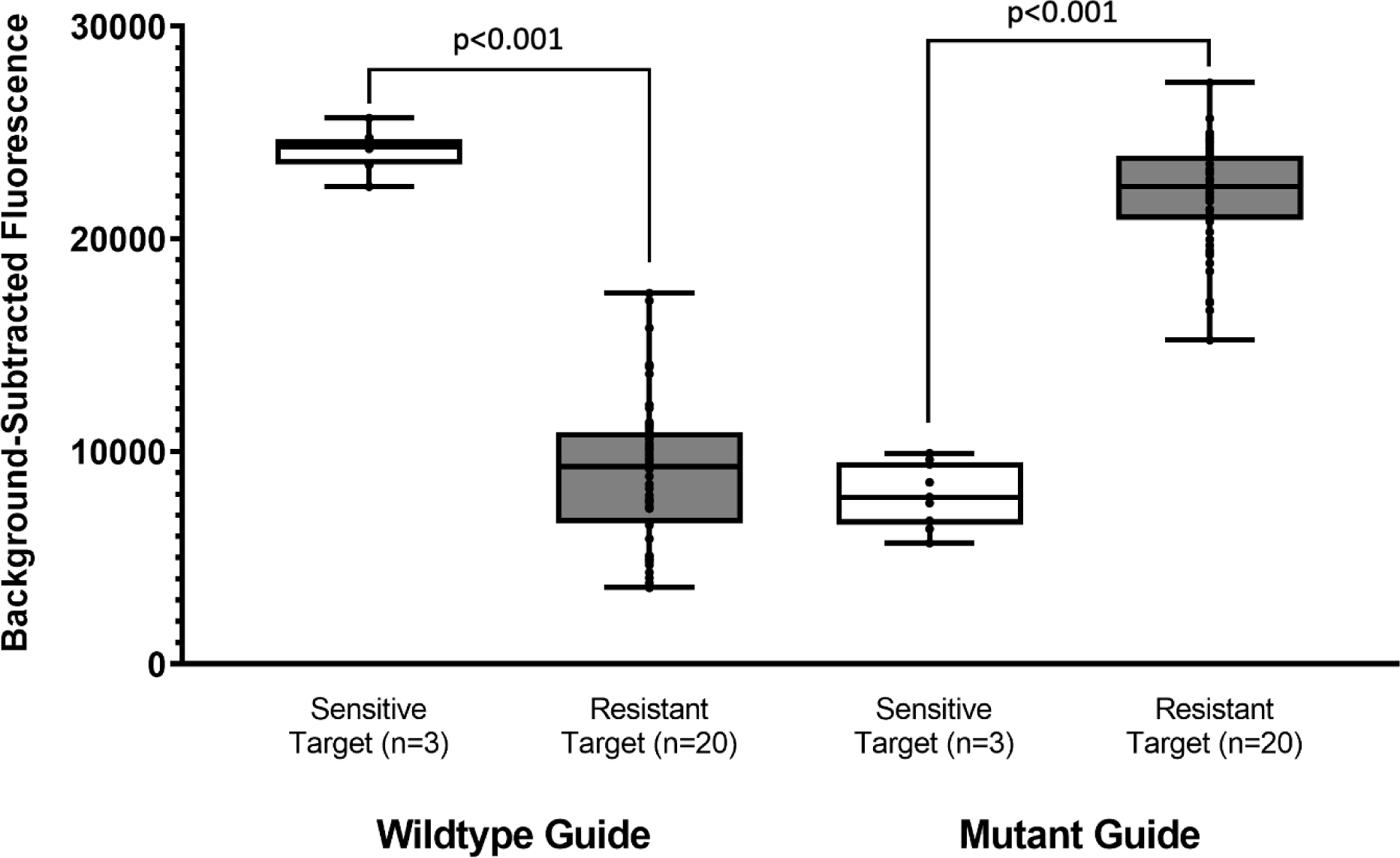
Cas13a-Based Gyrase A Determination of Purified *N. gonorrhoeae* Specimens. Figure 4 Legend: The figure shows the pooled discrimination of the Cas13a-based assay using fluorescence detection for determining the *gyr*A genotype of 23 purified *N. gonorrhoeae* isolates.

**Figure 5:**
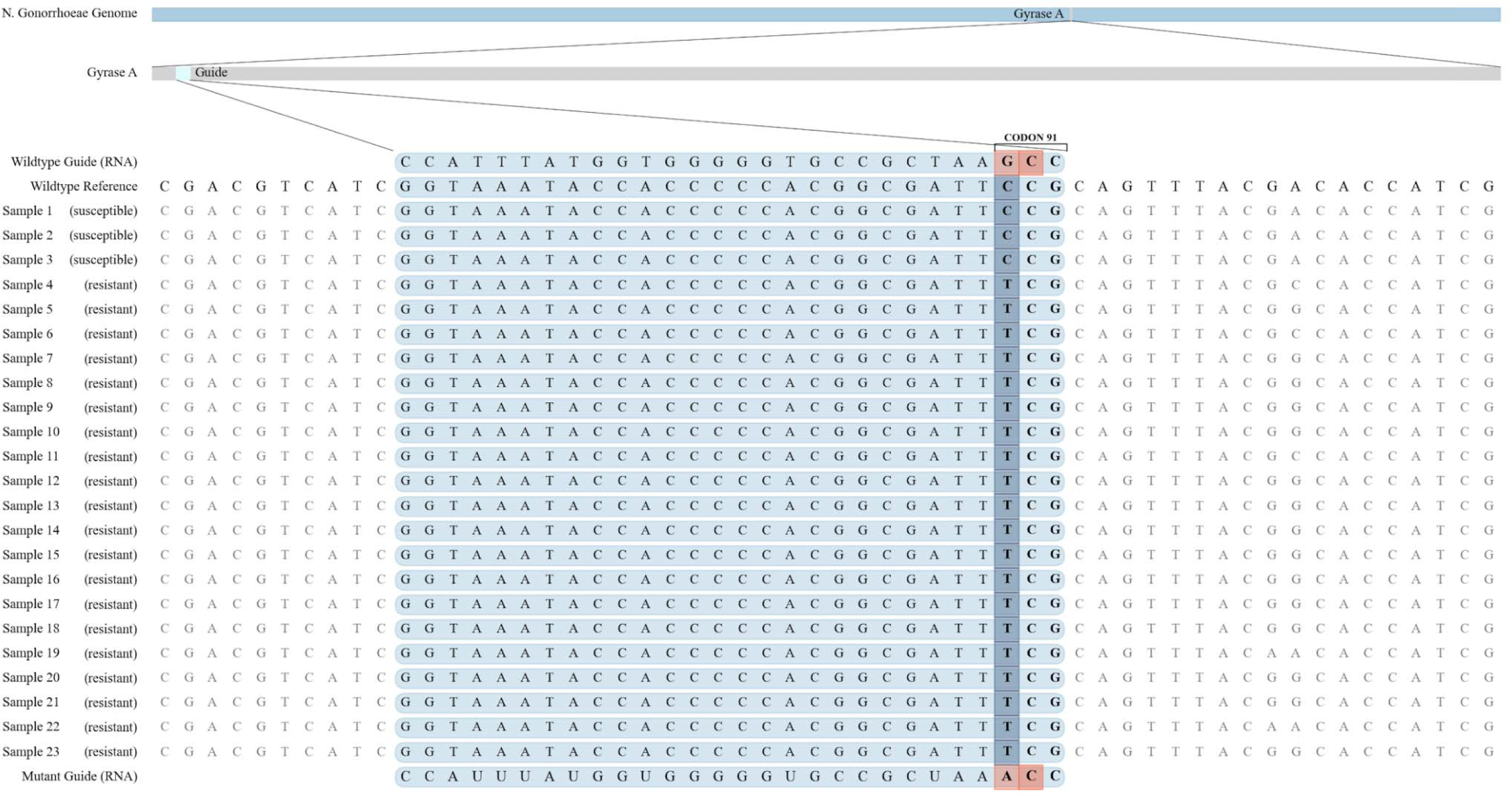
DNA Sequence Alignment of Codon 91 of the *gyr*A Gene from 23 Purified *N. gonorrhoeae* Isolates. Figure 5 Legend: The figure shows the DNA sequence alignment of codon 91 of the *gyr*A gene in *N. gonorrhoeae* with the two CRISPR-Cas13a guide sequences.

We next aimed to convert the *gyr*A resistance assay into a portable format suitable for use in resource-limited settings. We tested a lateral flow format, again substituting the standard fluorescence reporter with a biotinylated FAM reporter compatible with the test strips. Figure 6a shows the performance of the *gyr*A lateral flow on 3 purified isolates (1 with known phenotypic and genotypic susceptibility to ciprofloxacin and 2 with known resistance). We tested each isolate in duplicate. The wildtype guide failed to discriminate visually between resistant and susceptible isolates. The mutant guide demonstrated promising discrimination; however, we detected a faint positive line in the susceptible isolate.

**Figure 6:**
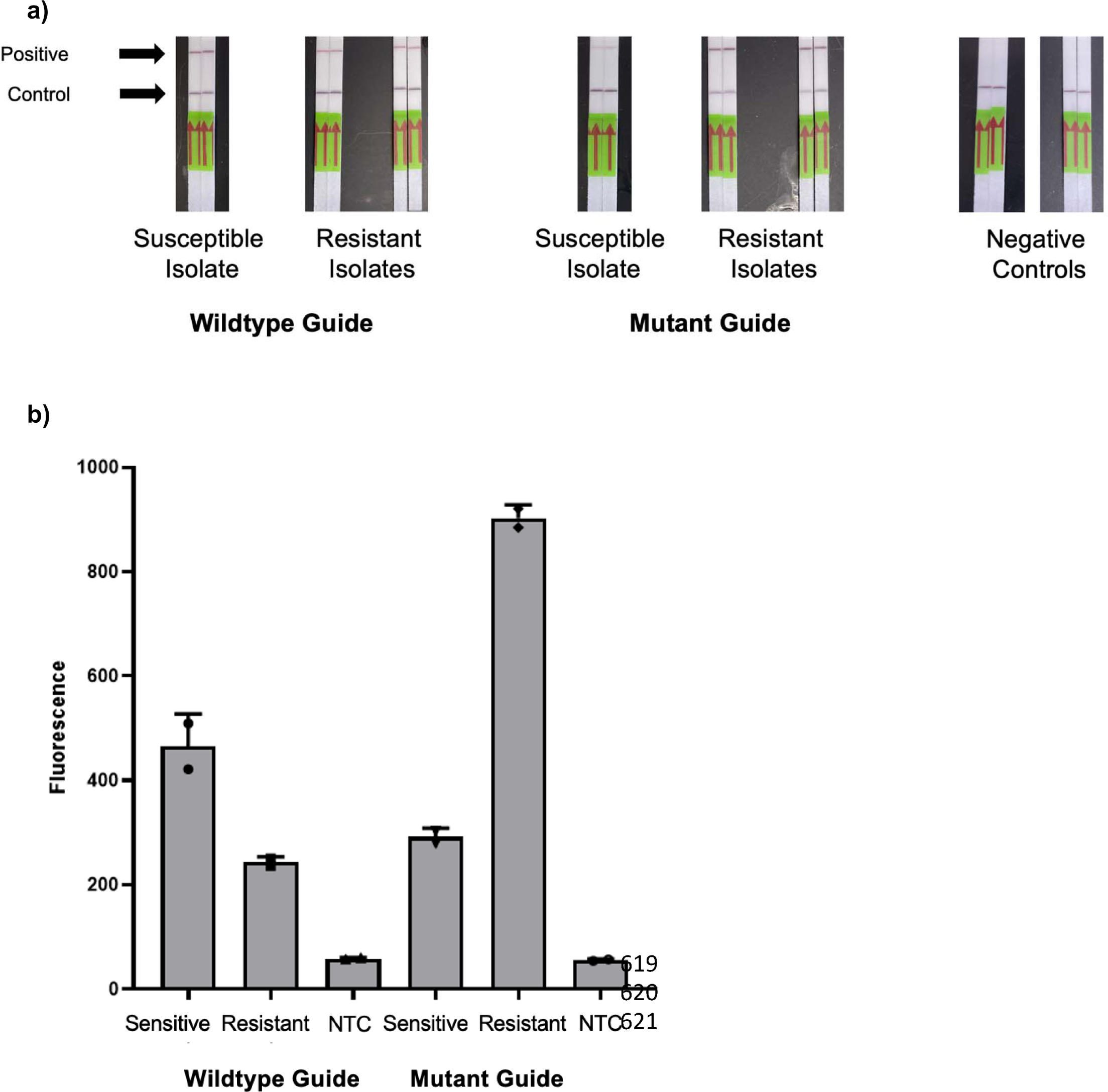
Performance of a Cas13a-Based *gyr*A Assay Using Lateral Flow Strips and a Portable Quantitative Fluorometer on Purified *N. gonorrhoeae* Isolates. Figure 6 Legend: Panel a) shows the performance of a Cas13a-based lateral flow assay using both wildtype and mutant guides for determining *gyr*A genotype among 3 *N. gonorrhoeae* isolates. Panel b) shows the same Cas13a assay read on a Qubit 4 Fluorometer. NTC (Negative Template Control)

Given the technical limitations of our *gyr*A assay using a lateral flow readout, we evaluated the performance of the assay using a portable quantitative fluorescence detector. Such a detector, the Qubit 4 Fluorometer (ThermoFisher Scientific, United States), would permit low-cost detection in the absence of a plate reader (Cytation 5, BioTek, United States). We incubated our one-pot SHERLOCK reaction for 90 minutes at 37° C and then transferred the reaction to Qbit Assay tubes, diluted with nuclease–free water to 200 µL. We measured green fluorescence detection on the blue excitation setting (430-495 excitation filter; 510-580 emission filter). Figure 6b shows successful discrimination for both the wildtype and mutant isolates using this method.

## Discussion

We report the development of a Cas13a-based lateral flow *N. gonorrhoeae* detection assay able to detect 100% of tested isolates and does not amplify closely-related *Neisseria* species. That assay offers the potential to introduce pathogen-specific diagnostics into low-resource settings that lack infrastructure for complex laboratory-based testing. More work is needed to establish the sensitivity and specificity of the assay in a clinical setting and to optimize its performance to meet World Health Organization standards for point-of-care tests (48). That includes the development of methods that could omit an extraction step and minimizing time to detection. Our preliminary results indicate that thermal extraction is a promising strategy. While 90 minutes was allocated for the lateral flow incubation to standardize our findings with prior protocols, peak fluorescence was noted at 20 minutes, indicating that the assay could provide rapid results in the field.

We also report the development of a Cas13a-based fluorescence detection assay with excellent discrimination of wildtype and mutant *gyr*A genotype isolates for predicting ciprofloxacin resistance. That assay showed a 100% agreement with both phenotypically and genotypically determined resistance to ciprofloxacin. Given the urgent need to combat antimicrobial-resistant *N, gonorrhoeae* infections (14, 15), and the high burden of resistance in resource-limited settings (31–33), such an assay may permit resistance-guided therapy without expensive laboratory equipment. While promising, the lateral flow Cas13a *gyr*A assay was not able to discriminate between wildtype and mutant genotypes as definitively, and will require further optimization. Iterative adaptations of guide sequences and position of the mutation of interest and the synthetic mutation relative to the Cas hairpin may improve the specificity of the assay on the lateral flow platform. Additional optimization will also be required to reduce the time involved in running the assay.

As an alternative field-deployable method for determining ciprofloxacin resistance, we devised a method for portable fluorescence of *gyr*A genotypes that overcame limitations of the lateral flow format for that assay. The Cas13a *gyr*A assay showed excellent discrimination between sensitive and resistant genotypes and can be implemented in resource-limited settings much more easily than qPCR or the BioTek Cytation 5 plate reader. While more expensive than paper-based assays and electricity-dependent, that fluorescence-based approach would still permit rapid and portable *gyr*A genotyping of *N. gonorrhoeae* specimens. With minor modifications, such as lyophilization of reagents and optimization of reaction conditions, we believe that some resource-constrained areas with basic laboratory infrastructure could consider assessing the feasibility of *N. gonorrhoeae* detection and *gyr*A genotyping using that assay format.

Our study had several important limitations. First, while we report on the *in vitro* performance of two newly described assays, our study evaluated the performance of those assays on a small number of isolates, thus limiting the precision of our findings. Moreover, the clinical utility remains to be determined and requires evaluation in a clinical setting. In particular, the processing required of those specimens will be of particular relevance for low-resource settings with limited laboratory infrastructure. However, while other rapid diagnostics for sexually transmitted infections are increasingly available (49), none has been sufficiently low-cost, timely, and user friendly to be optimally suited for low-resource settings, and none has incorporated detection of molecular markers of resistance. Thus, our results may provide the groundwork for introducing point-of-care resistance-guided therapy into settings previously constrained to syndromic management.

## Conclusion

We developed a paper-based lateral flow Cas13a assay for detecting *N. gonorrhoeae* which was able to detect *N. gonorrhoeae* and discriminate between other *Neisseria* species. We also developed a fluorescence-based Cas13a assay for determining *gyr*A genotype, which demonstrated excellent discrimination for both phenotypic and genotypic ciprofloxacin resistance among purified isolates.

## Data Availability

All data produced in the present study are available upon reasonable request to the authors

## Acknowledgements and Funding

This work was supported in part by the Massachusetts General Hospital Department of Medicine Innovation Program grant to R.H.G, NIH NIAID U19AI110818 to P.C.S, and grants 2019123 and 2021287 from the Doris Duke Charitable Foundation to J.E.L. The authors would like to acknowledge Kevin Ard and Jana Jarolimova for their support of this project as well as Benjamin Kotzen for his assistance with Figure 5.

## References

1. Kirkcaldy RD, Weston E, Segurado AC, Hughes G. Epidemiology of gonorrhoea: a global perspective. Sex Health. 2019;16(5):401–11.

2. Rowley J, Vander Hoorn S, Korenromp E, Low N, Unemo M, Abu-Raddad LJ, et al. Chlamydia, gonorrhoea, trichomoniasis and syphilis: global prevalence and incidence estimates, 2016. Bull World Health Organ. 2019;97(8):548-62P.

3. Vaezzadeh K, Sepidarkish M, Mollalo A, As’adi N, Rouholamin S, Rezaeinejad M, et al. Global prevalence of Neisseria gonorrhoeae infection in pregnant women: a systematic review and meta-analysis. Clin Microbiol Infect. 2023;29(1):22–31.

4. Whelan J, Abbing-Karahagopian V, Serino L, Unemo M. Gonorrhoea: a systematic review of prevalence reporting globally. BMC Infect Dis. 2021;21(1):1152.

5. Mitchell C, Prabhu M. Pelvic inflammatory disease: current concepts in pathogenesis, diagnosis and treatment. Infect Dis Clin North Am. 2013;27(4):793–809.

6. Tsevat DG, Wiesenfeld HC, Parks C, Peipert JF. Sexually transmitted diseases and infertility. Am J Obstet Gynecol. 2017;216(1):1–9.

7. Dolange V, Churchward CP, Christodoulides M, Snyder LAS. The Growing Threat of Gonococcal Blindness. Antibiotics (Basel). 2018;7(3).

8. Jarvis GA, Chang TL. Modulation of HIV transmission by Neisseria gonorrhoeae: molecular and immunological aspects. Curr HIV Res. 2012;10(3):211–7.

9. Galvin SR, Cohen MS. The role of sexually transmitted diseases in HIV transmission. Nat Rev Microbiol. 2004;2(1):33–42.

10. Chesson HW, Pinkerton SD. Sexually transmitted diseases and the increased risk for HIV transmission: implications for cost-effectiveness analyses of sexually transmitted disease prevention interventions. J Acquir Immune Defic Syndr. 2000;24(1):48–56.

11. Zetola NM, Bernstein KT, Wong E, Louie B, Klausner JD. Exploring the relationship between sexually transmitted diseases and HIV acquisition by using different study designs. J Acquir Immune Defic Syndr. 2009;50(5):546–51.

12. Cohen MS, Council OD, Chen JS. Sexually transmitted infections and HIV in the era of antiretroviral treatment and prevention: the biologic basis for epidemiologic synergy. J Int AIDS Soc. 2019;22 Suppl 6(Suppl Suppl 6):e25355.

13. Jones J, Weiss K, Mermin J, Dietz P, Rosenberg ES, Gift TL, et al. Proportion of Incident Human Immunodeficiency Virus Cases Among Men Who Have Sex With Men Attributable to Gonorrhea and Chlamydia: A Modeling Analysis. Sex Transm Dis. 2019;46(6):357–63.

14. United Nations News. Antimicrobial resistance a ‘global health emergency,’ UN, ahead of awareness week. Nov 12 2018. Available at: https://news.un.org/en/story/2018/11/1025511 Accessed January 11, 2023.

15. Centers for Disease, Control. Antibiotic resistance threats in the United States, 2013. Atlanta: CDC. 2013.

16. Unemo M, Shafer WM. Antimicrobial resistance in Neisseria gonorrhoeae in the 21st century: past, evolution, and future. Clin Microbiol Rev. 2014;27(3):587–613.

17. Ventola CL. The antibiotic resistance crisis: part 1: causes and threats. P T. 2015;40(4):277–83.

18. Kueakulpattana N, Wannigama DL, Luk-In S, Hongsing P, Hurst C, Badavath VN, et al. Multidrug-resistant Neisseria gonorrhoeae infection in heterosexual men with reduced susceptibility to ceftriaxone, first report in Thailand. Sci Rep. 2021;11(1):21659.

19. Bala M, Sood S. Cephalosporin Resistance in Neisseria gonorrhoeae. J Glob Infect Dis. 2010;2(3):284–90.

20. Gianecini R, Oviedo C, Stafforini G, Galarza P. Neisseria gonorrhoeae Resistant to Ceftriaxone and Cefixime, Argentina. Emerg Infect Dis. 2016;22(6):1139–41.

21. Unemo M, Golparian D, Nicholas R, Ohnishi M, Gallay A, Sednaoui P. High-level cefixime– and ceftriaxone-resistant Neisseria gonorrhoeae in France: novel penA mosaic allele in a successful international clone causes treatment failure. Antimicrob Agents Chemother. 2012;56(3):1273–80.

22. Wi T, Lahra MM, Ndowa F, Bala M, Dillon JR, Ramon-Pardo P, et al. Antimicrobial resistance in Neisseria gonorrhoeae: Global surveillance and a call for international collaborative action. PLoS Med. 2017;14(7):e1002344.

23. Workowski KA, Bachmann LH, Chan PA, Johnston CM, Muzny CA, Park I, et al. Sexually Transmitted Infections Treatment Guidelines, 2021. MMWR Recomm Rep. 2021;70(4):1-187.

24. Buono SA, Watson TD, Borenstein LA, Klausner JD, Pandori MW, Godwin HA. Stemming the tide of drug-resistant Neisseria gonorrhoeae: the need for an individualized approach to treatment. J Antimicrob Chemother. 2015;70(2):374–81.

25. Tuite AR, Gift TL, Chesson HW, Hsu K, Salomon JA, Grad YH. Impact of Rapid Susceptibility Testing and Antibiotic Selection Strategy on the Emergence and Spread of Antibiotic Resistance in Gonorrhea. J Infect Dis. 2017;216(9):1141–9.

26. Schink JC, Keith LG. Problems in the culture diagnosis of gonorrhea. J Reprod Med. 1985;30(3 Suppl):244-9.

27. Otieno FO, Ndivo R, Oswago S, Ondiek J, Pals S, McLellan-Lemal E, et al. Evaluation of syndromic management of sexually transmitted infections within the Kisumu Incidence Cohort Study. Int J STD AIDS. 2014;25(12):851–9.

28. Verwijs MC, Agaba SK, Sumanyi JC, Umulisa MM, Mwambarangwe L, Musengamana V, et al. Targeted point-of-care testing compared with syndromic management of urogenital infections in women (WISH): a cross-sectional screening and diagnostic accuracy study. Lancet Infect Dis. 2019;19(6):658–69.

29. Portnoy J, Mendelson J, Clecner B, Heisler L. Asymptomatic gonorrhea in the male. Can Med Assoc J. 1974;110(2):169 passim.

30. Handsfield HH, Lipman TO, Harnisch JP, Tronca E, Holmes KK. Asymptomatic gonorrhea in men. Diagnosis, natural course, prevalence and significance. N Engl J Med. 1974;290(3):117–23.

31. Iwuji C, Pillay D, Shamu P, Murire M, Nzenze S, Cox LA, et al. A systematic review of antimicrobial resistance in Neisseria gonorrhoeae and Mycoplasma genitalium in sub-Saharan Africa. J Antimicrob Chemother. 2022;77(8):2074–93.

32. Kakooza F, Musinguzi P, Workneh M, Walwema R, Kyambadde P, Mande E, et al. Implementation of a standardised and quality-assured enhanced gonococcal antimicrobial surveillance programme in accordance with WHO protocols in Kampala, Uganda. Sex Transm Infect. 2021;97(4):312–6.

33. Crucitti T, Belinga S, Fonkoua MC, Abanda M, Mbanzouen W, Sokeng E, et al. Sharp increase in ciprofloxacin resistance of Neisseria gonorrhoeae in Yaounde, Cameroon: analyses of a laboratory database period 2012-2018. Int J STD AIDS. 2020;31(6):579-86.

34. Global action plan to control the spread and impact of antimicrobial resistance in Neisseria gonorrhoeae. Geneva: WHO; 2012.

35. Whiley DM, Anderson TP, Barratt K, Beaman MH, Buda PJ, Carter M, et al. Evidence that the gonococcal porA pseudogene is present in a broad range of Neisseria gonorrhoeae strains; suitability as a diagnostic target. Pathology. 2006;38(5):445–8.

36. Allan-Blitz LT, Wang X, Klausner JD. Wild-Type Gyrase A Genotype of Neisseria gonorrhoeae Predicts In Vitro Susceptibility to Ciprofloxacin: A Systematic Review of the Literature and Meta-Analysis. Sex Transm Dis. 2017;44(5):261–5.

37. Allan-Blitz LT, Adamson PC, Klausner JD. Resistance-Guided Therapy for Neisseria gonorrhoeae. Clin Infect Dis. 2022;75(9):1655–60.

38. Gootenberg JS, Abudayyeh OO, Lee JW, Essletzbichler P, Dy AJ, Joung J, et al. Nucleic acid detection with CRISPR-Cas13a/C2c2. Science. 2017;356(6336):438-42.

39. Lobato IM, O’Sullivan CK. Recombinase polymerase amplification: Basics, applications and recent advances. Trends Analyt Chem. 2018;98:19–35.

40. Myhrvold C, Freije CA, Gootenberg JS, Abudayyeh OO, Metsky HC, Durbin AF, et al. Field-deployable viral diagnostics using CRISPR-Cas13. Science. 2018;360(6387):444-8.

41. de Puig H, Lee RA, Najjar D, Tan X, Soeknsen LR, Angenent-Mari NM, et al. Minimally instrumented SHERLOCK (miSHERLOCK) for CRISPR-based point-of-care diagnosis of SARS-CoV-2 and emerging variants. Sci Adv. 2021;7(32).

42. Abudayyeh OO, Gootenberg JS, Konermann S, Joung J, Slaymaker IM, Cox DB, et al. C2c2 is a single-component programmable RNA-guided RNA-targeting CRISPR effector. Science. 2016;353(6299):aaf5573.

43. Joung J, Ladha A, Saito M, Segel M, Bruneau R, Huang MW, et al. Point-of-care testing for COVID-19 using SHERLOCK diagnostics. medRxiv. 2020.

44. Buckley C, Trembizki E, Donovan B, Chen M, Freeman K, Guy R, et al. A real-time PCR assay for direct characterization of the Neisseria gonorrhoeae GyrA 91 locus associated with ciprofloxacin susceptibility. J Antimicrob Chemother. 2016;71(2):353–6.

45. Heiniger EK, Buser JR, Mireles L, Zhang X, Ladd PD, Lutz BR, et al. Comparison of point– of-care-compatible lysis methods for bacteria and viruses. J Microbiol Methods. 2016;128:80–7.

46. Metsky HC, Welch NL, Pillai PP, Haradhvala NJ, Rumker L, Mantena S, et al. Designing sensitive viral diagnostics with machine learning. Nat Biotechnol. 2022;40(7):1123–31.

47. Kellner MJ, Koob JG, Gootenberg JS, Abudayyeh OO, Zhang F. SHERLOCK: nucleic acid detection with CRISPR nucleases. Nat Protoc. 2019;14(10):2986–3012.

48. Land KJ, Boeras DI, Chen XS, Ramsay AR, Peeling RW. REASSURED diagnostics to inform disease control strategies, strengthen health systems and improve patient outcomes. Nat Microbiol. 2019;4(1):46–54.

49. Adamson PC, Loeffelholz MJ, Klausner JD. Point-of-Care Testing for Sexually Transmitted Infections: A Review of Recent Developments. Arch Pathol Lab Med. 2020;144(11):1344–51.

