## Supplement for "Development of Cas13a-based Assays for *Neisseria gonorrhoeae* Detection and Gyrase A Determination"

**Supplemental Table 1: Reagents and stock concentrations used in the development of the Cas13a-based *Neisseria gonorrhoeae* and *gyr*A assays, qPCR, DNA Sequencing, and Lateral Flow.**

| **Reagent** | **Reaction** | **Source** | **Stock Concentration** | **Notes** |
| --- | --- | --- | --- | --- |
| C2c2 *Lwa*Cas13a | SHERLOCK | GenScript | 5 mg/mL |  |
| Rnase Inhibitor | SHERLOCK | NEB | 40 U/µL | Murine |
| T7 RNA Polymerase | SHERLOCK | Lucigen | 50 U/µL | NextGen |
| Reaction Buffer | SHERLOCK | N/A | 5X | 0.1 M HEPES pH 8.0; 300 mM KCl; 25% PEG-8000 |
| rNTPs | SHERLOCK | NEB | 25 mM of each nucleotide |  |
| RNase Alert Substrate v2 | SHERLOCK | Thermo Fisher Scientific | 2 µM |  |
| MgAc | SHERLOCK | TwistDx | 280 mM | TwistAmp Basic Kit |
| Storage Buffer | SHERLOCK | N/A | 1X | 50 mM Tris pH 7.5; 600 mM KCl; 5% glycerol; 2 mM DTT |
| RPA Primers  (forward, reverse) | SHERLOCK | Integrated DNA Technologies | 50 µM of each primer | Supplementary Table 2 for sequences |
| Cas13a crRNA  (wildtype, mutant, por A) | SHERLOCK | Integrated DNA Technologies | 2.5 µM of each | Supplementary Table 2 for sequences |
| Nuclease-Free H2O |  | Thermo Fisher Scientific | N/A | Invitrogen |
| RPA Pellets (lyophilized) | SHERLOCK | TwistDx | N/A | TwistAmp Basic Kit |
| Synthetic DNA Target (wildtype, mutant) | SHERLOCK | Integrated DNA Technologies | 10^10^ copies/µL | Supplementary Table 2 for sequences |
| PCR Primers (forward, reverse) | qPCR | Integrated DNA Technologies | 100 µM of each primer | Supplementary Table 2 for sequences |
| FastStart SYBR Green Master | qPCR | Roche | 2X |  |
| Lateral Flow Dipsticks | SHERLOCK | Milenia Biotec | N/A | HybriDetect - Universal Lateral Flow Assay Kit |
| HybriDetect Assay Buffer | SHERLOCK | Milenia Biotec | N/A | HybriDetect - Universal Lateral Flow Assay Kit |

**Supplementary Table 2: Primer, Guide, and Target sequences for qPCR and Cas13a-based assays.**

| **Reagent** | **Sequence** |
| --- | --- |
| Por A RPA Primer 1.1 (forward) | GAAATTAATACGACTCACTATAGGGTGTATTATGCCGGTCTGAATTACAAAAATG |
| Por A RPA Primer 1.1 (reverse) | GTACCTGATGGTTTTTCAATGGATCGGTATC |
| Por A RPA Primer 1.2 (forward) | GAAATTAATACGACTCACTATAGGTGTGTATTATGCCGGTCTGAATTACAAAAATG |
| Por A RPA Primer 1.2 (reverse) | TACCTGATGGTTTTTCAATGGATCGGTATCAC |
| Por A RPA Primer 2.1 (forward) | GAAATTAATACGACTCACTATAGGCATCAGCTATGCCCATGGTTTCGACTTTGTC |
| Por A RPA Primer 2.1 (reverse) | GAAGTGCGCTTGGAAAAATCGTAATCGACAC |
| Por A RPA Primer 2.2 (forward) | GAAATTAATACGACTCACTATAGGCACTGCTTCCTACCGCTTCGGTAATACAGTC |
| Por A RPA Primer 2.2 (reverse) | GAAGTGCGCTTGGAAAAATCGTAATCGACACC |
| Por A RPA Primer 3.1 (forward) | GAAATTAATACGACTCACTATAGGAATAATAATGTGGCTTCGCAATTGGGTATTTT |
| Por A RPA Primer 3.1 (reverse) | CATATTTAAGGGCATAATTTCCGAAAAAGC |
| Por A RPA Primer 3.2 (forward) | GAAATTAATACGACTCACTATAGGATAATAATGTGGCTTCGCAATTGGGTATTTTC |
| Por A RPA Primer 3.2 (reverse) | GCATATTTAAGGGCATAATTTCCGAAAAAG |
| Por A Cas 13a crRNA 1 | GAUUUAGACUACCCCAAAAACGAAGGGGACUAAAACUCGCAUAUUUAAGGGCAUAAUUUCCGAA |
| Por A Cas 13a crRNA 2 | GAUUUAGACUACCCCAAAAACGAAGGGGACUAAAACGAUUUGAUCAUAGCUGGUAUGUUCGCGU |
| Por A Cas 13a crRNA 3 | GAUUUAGACUACCCCAAAAACGAAGGGGACUAAAACAUAGGCGGACUUGCUGUUUUGACUCGGA |
| Gyrase A RPA Primer 1 (forward) | GAAATTAATACGACTCACTATAGGACTGGAATG CCGCCTACAAA |
| Gyrase A RPA Primer 1 (reverse) | CCCTGTCCGTCTATCAGCAC |
| Gyrase A RPA Primer 2 (forward) | GAAATTAATACGACTCACTATAGGAACTGGAATGCC GCCTACAA |
| Gyrase A RPA Primer 2 (reverse) | GTTGCCCTGTCCGTCTATCA |
| Gyrase A RPA Primer 3 (forward) | GAAATTAATACGACTCACTATAGGATAACTGGAATG CCGCCTACA |
| Gyrase A RPA Primer 3 (reverse) | TTTTGCGCCATACGGACGAT |
| Gyrase A Cas13a crRNA 1 (wildtype) | GAUUUAGACUACCCCAAAAACGAAGGGGACUAAAACCCGAAUCGCCGUGGGGGUGGUAUUUACC |
| Gyrase A Cas13a crRNA 1 (mutant) | GAUUUAGACUACCCCAAAAACGAAGGGGACUAAAACCCAAAUCGCCGUGGGGGUGGUAUUUACC |
| Gyrase A Cas13a crRNA 2 (wildtype) | GAUUUAGACUACCCCAAAAACGAAGGGGACUAAAACCGGUAUCGCCGUGGGGGUGGUAUUUACC |
| Gyrase A Cas13a crRNA 2 (mutant) | GAUUUAGACUACCCCAAAAACGAAGGGGACUAAAACCGAUAUCGCCGUGGGGGUGGUAUUUACC |
| Gyrase A Synthetic DNA Target (wildtype) | GGTAAATACCACCCCCACGGCGATTCCG |
| Gyrase A Synthetic DNA Target (mutant) | GGTAAATACCACCCCCACGGCGATTTCG |
| T7 promoter | GAAATTAATACGACTCACTATAGG |
| Gyrase A PCR Primer (forward) | GCGACGGCCTAAAGCCAGTG |
| Gyrase A PCR Primer (reverse) | GTCTGCCAGCATTTCATGTGAG |

**Supplemental Figure 1: Guide RNA Performance for detecting synthetic DNA with mutant and wildtype *gyr*A genotypes via a Cas13a-based detection system**

**a)	b)**

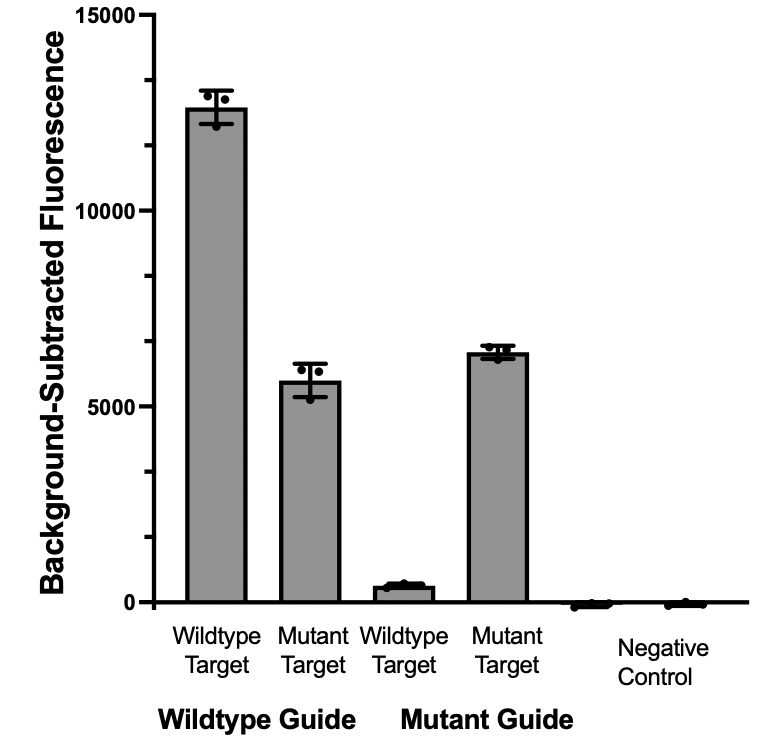

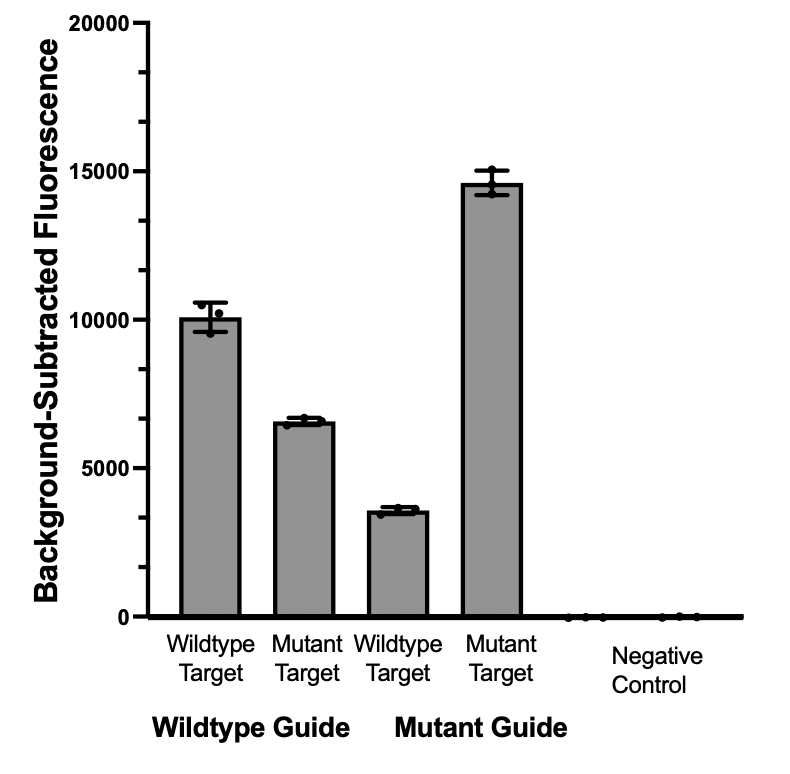

**c)**
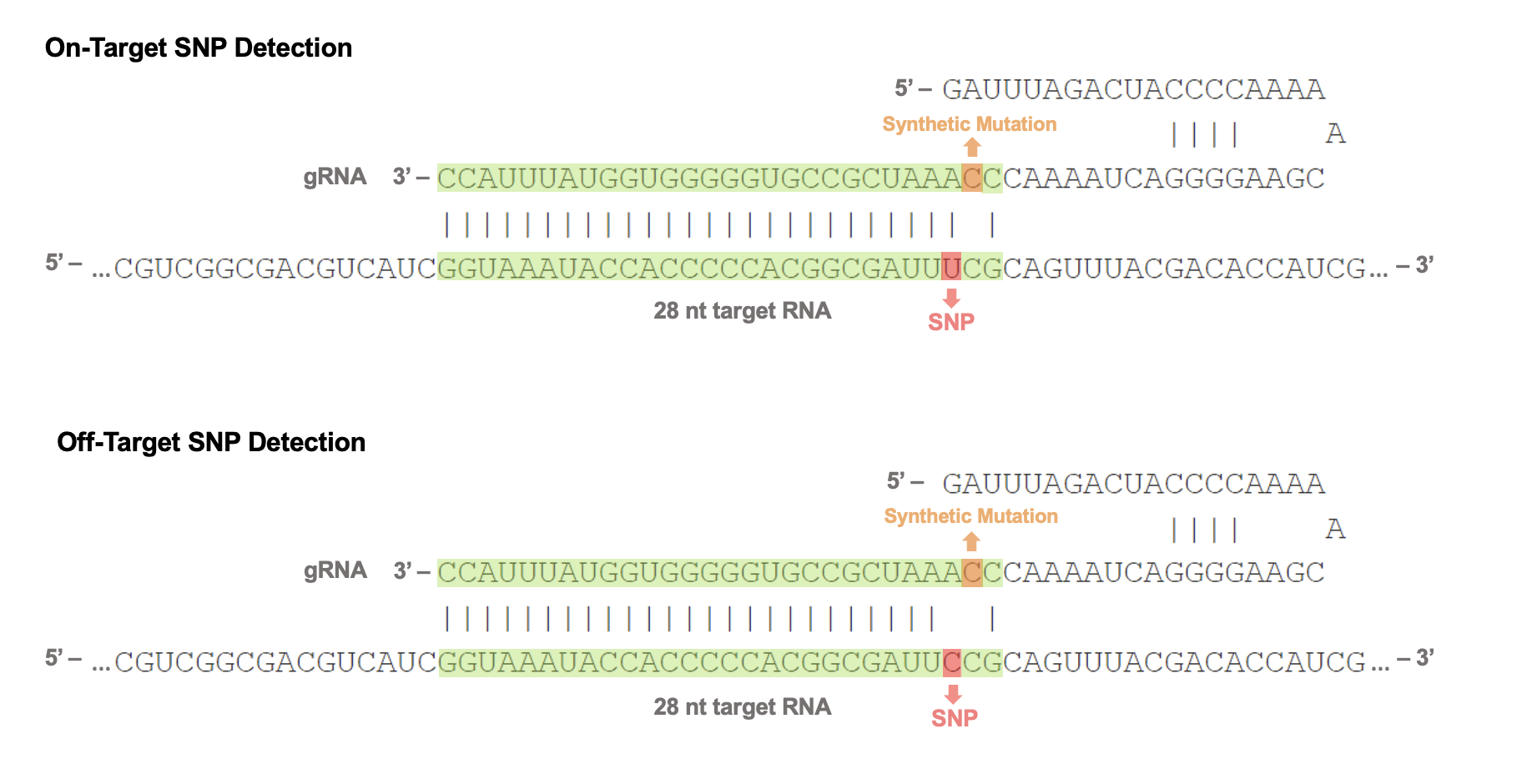

Supplemental Figure 1 Legend: Panel a) shows the discrimination between wildtype and mutant *gyr*A DNA of the gRNA with the synthetic mutation at the second position from the Cas13a enzyme; Panel b) shows the discrimination between wildtype and mutant *gyr*A DNA of the gRNA with the synthetic mutation at the fourth position from the Cas13a enzyme; Panel c) shows the selected gRNA sequence and alignment.

**Supplemental Figure 2: Performance of three primer sets for amplification of synthetic DNA with mutant and wildtype *gyr*A genotypes**

**a)**	**b)**

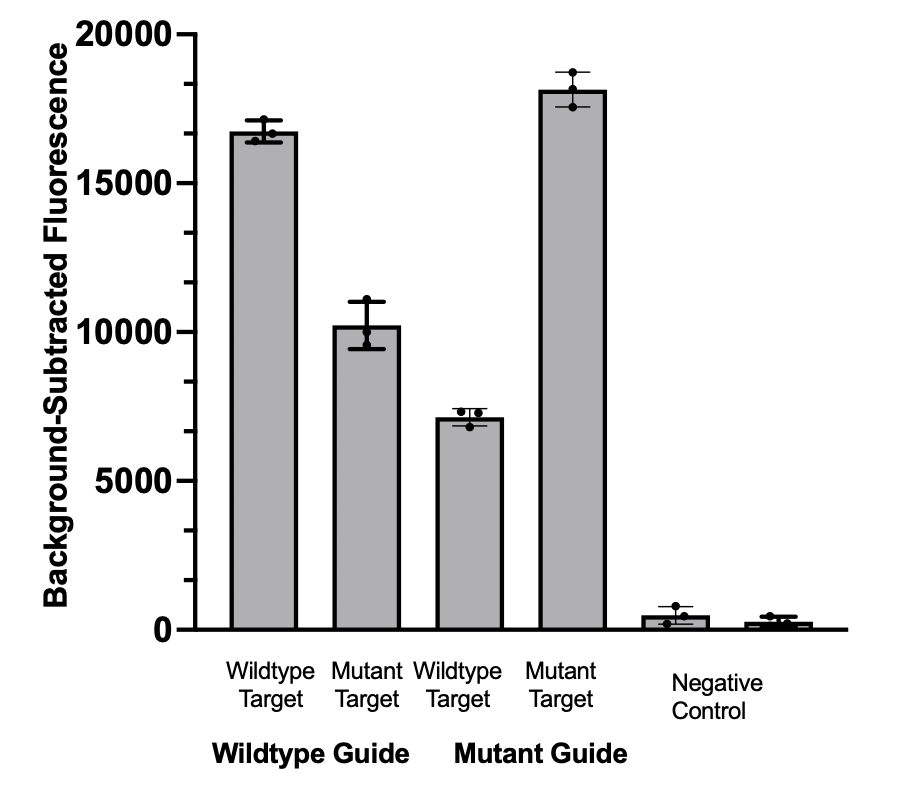

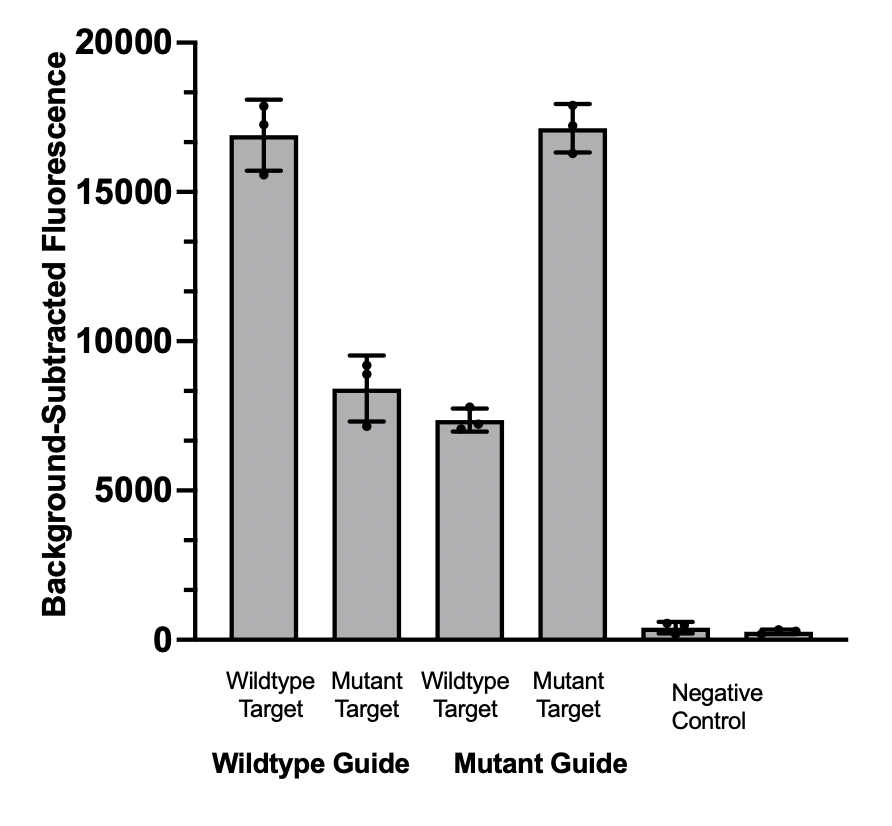

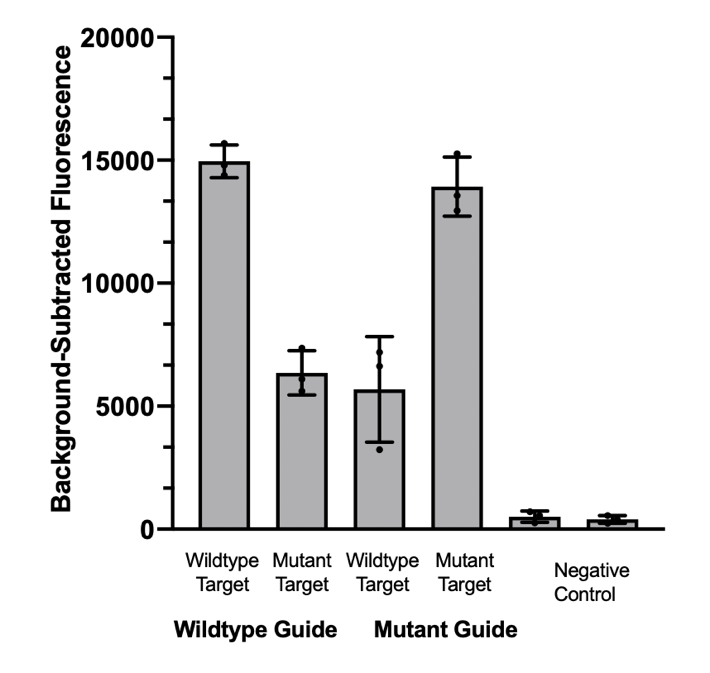
**c)**

Supplemental Figure 2 Legend: The figure shows the performance of three different primer sets using the selected gRNA in amplifying the synthetic wildtype and mutant *gyr*A DNA sequences. The primer set in panel a) was selected as it demonstrated the highest amplification while preserving discrimination with the selected gRNA.

**Supplemental Figure 3: Phylogenetic Tree of 23 *N. gonorrhoeae* isolates aligned with reference *N. gonorrhoeae genome***

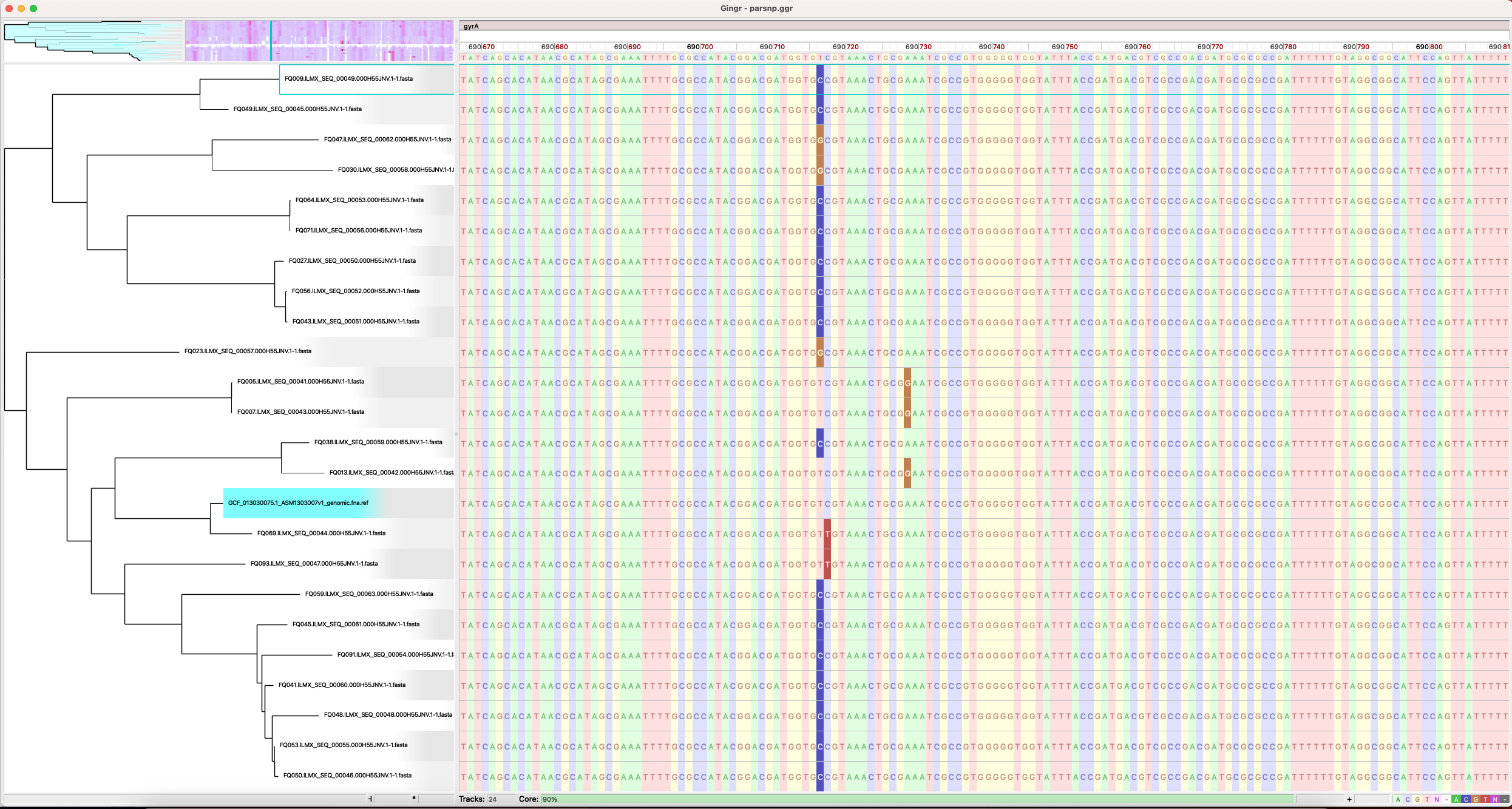

**Supplemental Figure 4: Discrimination of the Cas13a-based assay among 23 purified *Neisseria gonorrhoeae* isolates.**

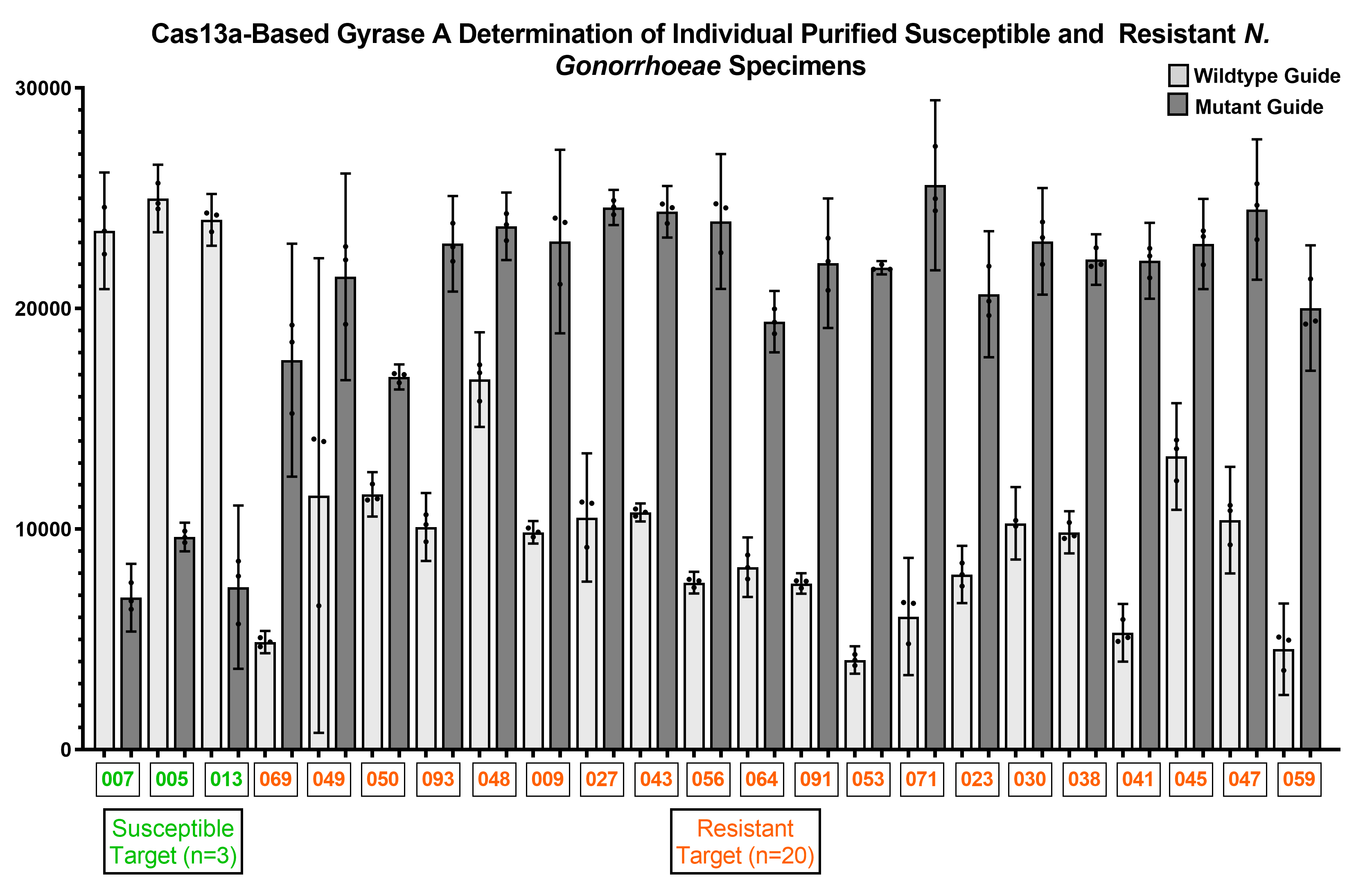
